# Sociodemographic and clinical risk factors for suicidal ideation and suicide attempt in functional/dissociative seizures and epilepsy: a large cohort study

**DOI:** 10.1101/2023.11.27.23298811

**Authors:** Irene Faiman, John Hodsoll, Iman Jasani, Allan H. Young, Paul Shotbolt

## Abstract

**Objective:** To identify risk factors for first episodes of suicidal ideation and suicide attempt in people with functional/dissociative seizures (FDS) or epilepsy.

**Methods:** Retrospective cohort study from the UK’s largest tertiary mental health care provider, with linked national admission data from the Hospital Episode Statistics (HES). Participants were 2383 people with a primary or secondary diagnosis of FDS or epilepsy attending between 01/01/2007 and 18/06/2021. Outcomes were a first report of suicidal ideation and a first hospital admission for suicide attempt (ICD-10 X60-X84). Demographic and clinical risk factors were assessed using multivariable bias-reduced binomial-response generalised linear models.

**Results:** In both groups, ethnic minorities had significantly reduced odds of hospitalisations following a suicide attempt (OR: 0.45 – 0.49). Disorder-specific risk factors were gender, age, and comorbidity profile. In FDS, both genders had similar risk of suicidality; younger age was a risk factor for both outcomes (OR: 0.16 – 1.91) and a diagnosis of Depression or Personality Disorders was associated with higher odds of reporting suicidal ideation (OR: 1.91 and 3.01 respectively). In epilepsy, females had higher odds of being hospitalised following suicide attempt (OR: 1.64). Age had a quadratic association with both outcomes (OR: 0.88 – 1.06). A Substance Abuse Disorder was associated to higher suicidal ideation (OR: 2.67) whilst Developmental Disorders lowered the risk (OR: 0.16 – 0.24).

**Conclusions:** This is the first study systematically reporting risk factors for suicidality in people with FDS. Results for the large epilepsy cohort complement previous studies and will be useful in future meta-analyses.

**KEY MESSAGE BOX:**

**What is already known on this topic**

- People with epilepsy and people with functional/dissociative seizures (FDS) are at elevated risk of suicide
Identification of risk factors for suicidal ideation and suicide attempt in high-risk groups is essential to inform risk prevention strategies
**What this study adds**

- Several factors are associated with suicidality in FDS, including age, ethnicity, comorbid Depression and Personality Disorders.
- In epilepsy, suicidality is associated with age, gender, ethnicity, comorbid Substance Misuse Disorder and Developmental Disorders.
**How this study might affect research, practice or policy**

- Whilst disorder-specific factors will be useful to identify groups at higher risk in clinical settings, general risk factors can be target of population-based preventive strategies.

## INTRODUCTION

Suicide is one of the leading causes of death worldwide, and its prevention has been identified as a ‘global imperative’[1]. In the UK, the *Five Year Forward View for Mental Health* (2016) declared a national ambition of reducing suicide deaths by 10% by 2020. Whist national plans have been implemented to advance equality, coverage, and access to mental health services, the clinical and academic community should lead in identifying high-risk groups for improved clinical risk assessment and targeted preventive programs.

Suicidal ideation (thinking about, considering or planning suicide) and suicide attempts (self-harm with intent to die) are strong predictive factors of suicide death[2]. These immediate precursors of suicide can themselves lead to devastating consequences such as psychological burden, loss of employment, serious injury and disability. As certain factors might have greater or lesser influence in different clinical populations, there is urgent need to understand the risk factors for suicidal ideation and suicide attempts (collectively referred to as “suicidality” hereafter) to recognise people at risk and inform public health strategies.

To address this, the present study focuses on identifying factors associated with suicidality in two groups at elevated suicide risk: people with functional/dissociative seizures (FDS), and people with epilepsy. Epilepsy is a common neurological disorder characterised by an enduring propensity to experience epileptic seizures, i.e., transient signs and/or symptoms of excessive or synchronous neuronal activity[3]. In people with epilepsy, suicide rates are three times those observed in the general population[4]. FDS are abrupt and observable episodes of altered behaviour or consciousness that superficially resemble epileptic seizures but are not associated with electroencephalography changes[5]. People with FDS have elevated suicide mortality rates (2.6-18.8%) as compared to the general population[6–8], and the presence of FDS has been found to increase the risk of suicide attempt-related hospitalisation in people with epilepsy[7].

Several risk factors for suicidality in epilepsy have been identified[9–12]; we aim to provide additional evidence from a large cohort for the benefit of future aggregative studies like meta-analyses. On the other hand, evidence on the correlates of suicidality in FDS is scarce, with non-systematic reports from small samples[13,14]. To the best of our knowledge, this is the first study whose primary aim is to systematically investigate the sociodemographic and clinical factors associated with suicidal ideation and suicide attempt-related hospitalisation in this high-risk population.

## METHODS

### Design and setting

This is a retrospective cohort study. Data were extracted by a research nurse using the Clinical Record Interactive Search (CRIS)[15], a system providing de-identified information from South London and Maudsley NHS Foundation Trust (SLaM) electronic heath records. SLaM is the largest tertiary mental health care provider in Europe, with a catchment of four boroughs in southeast London with a population of over 1.2 million people, hosting the only NHS national specialist tertiary service in the UK offering assessment and treatment of severe dissociative disorders, including FDS.

Ethics approval for secondary analysis of CRIS data was obtained following the CRIS Oversight Committee’s review (project number 19-088; 14/11/2019 approval under ‘Oxfordshire C’ Research Ethics 23/SC/0257).

### Study population

The study’s source population were people attending SLaM between 01/01/2007 and 18/06/2021. Inclusion criteria were a primary or secondary diagnosis of epilepsy (International Classification of Diseases, version 10 (ICD-10) code G40) or FDS (ICD-10 F44.5). We also extracted data for people with concurrent diagnosis of epilepsy and FDS, which were utilised in a previous study based on the same data[7]; however, the small sample size prevented us from investigating risk and protective factors for this group. Exclusion criteria were a primary or secondary diagnosis of psychotic disorder (ICD-10 F20-F29) or structural brain disease, including cerebral malignancy (ICD-10 C71, C79.3, D43.0-D43.2, D49.6), traumatic brain injury (ICD-10 S01, S02, S06, S07, S09), dementia or progressive neurodegenerative disease (ICD-10 F00-F03).

### Outcome variables – Suicidal ideation and suicide attempt

The study outcomes were a first report of suicidal ideation and a first suicide attempt-related hospitalisation. Information on suicidal ideation was extracted using a Natural Language Processing (NLP) app, using Generalised Architecture for Text Engineering (GATE) and TextHunter software[16]. This has 87.8% precision and 91.7% recall to an instance of ideation[16]. To measure recall of the NLP app specifically for our sample, two researchers (I.F., I.J.) independently reviewed all 616 text snippets resulting in a positive ideation instance. It was coded whether this was a true and present instance (i.e., active within one year before the snippet date) or a true past instance (i.e., ever before the snippet date). Any disagreements were resolved upon discussion. Recall for present and past instances was 60% and 71% respectively. Inter-rater agreement was almost perfect (95.3%, Cohen’s K = 0.90). False positives were corrected to achieve higher accuracy in subsequent analyses.

Information on suicide attempt-related admissions was extracted from Hospital Episode Statistics (HES) admission data linked to our cohort. HES collates national-level data routinely collected by NHS providers (http://www.hscic.gov.uk/hes). People with a first UK registered hospital admission following suicide attempt (ICD-10 X60-X84) were considered cases of suicide attempt. Due to CRIS data access permissions, HES data were not extracted for patients under 18 at the time of data collection, and for those that were only ever seen in SLaM Hospital NHS Foundation Trust whilst under the age of 18.

### Sociodemographic and clinical variables

The following information was extracted: age, gender, ethnicity, and presence of comorbid neuropsychiatric diagnoses. Age at suicidal ideation and at suicide attempt was calculated; in the absence of suicidality, the age reported was the age at data extraction (18/06/2021), or age at death if deceased. Ethnicity was subdivided into White and other ethnicity (including Asian, Black, Mixed, Multiple and Other ethnicity). People were coded as having any neuropsychiatric diagnosis if at least one of the following disorders was recorded before the date of suicidal ideation or suicide attempt (or at any time if suicidality was not observed): Substance Misuse Disorders (ICD-10 F10-F19), Anxiety and stress-related disorders (including Obsessive Compulsive Disorder and Post Traumatic Stress Disorder; ICD-10 F40– F42, F43.1), Bipolar Disorder (ICD-10 F31) or Depressive Disorders (ICD-10 F32-F33), Personality Disorder (ICD-10 F60), Pervasive Developmental Disorders or moderate to profound Intellectual Disability (ICD-10 F71-F73, F84).

### Statistical analyses

We explored the association between sociodemographic and clinical factors and study outcomes separately for people with FDS and epilepsy. People were included independently of the diagnosis date. Our sample therefore includes people with a “lifetime diagnosis”, under the assumption that both disorders are associated with a neurobiological and neuropsychiatric phenotype existing even before seizure expression and diagnosis. To obtain an estimate of the total effect for each study variable, we first created causal diagrams describing the relationship between variables and outcomes, as recommended[17] (Supplementary Figures 1-4). These were built based on the clinical and academic expertise of a Consultant Neuropsychiatrist and a Consultant Psychiatrist (P.S. and A.Y.). The diagrams were then used to guide variable entry in multivariable bias reduced binomial-response generalised linear models[18] using the D3 likelihood ratio test for model comparison with multiply imputed data[19]. Age was assessed for non-linearity using restricted cubic splines[20], where models with zero (linear) to 5 knots were compared in terms of Akaike Information Criterion (AIC). Lowest AIC were for 3 knots in epilepsy and 4 in FDS (locations indicated in the regression tables). Covariates for each model are specified as footnotes to the result tables. As a rule of thumb, analyses were only performed for variables having at least 5 supporting observations per cell in cross-tabulation with the outcome[21]. Odds Ratios and associated Confidence Intervals were derived as a measure of association. Tjur’s coefficient of determination (pseudo-R^2^) was used to quantify the amount of shared variance between variables and outcomes[22]. Significance level was 0.05. Data were analysed using R statistical software[23] version 4.2, with the ‘brglm’ (0.72)[24] and ‘mice’ (3.15) package[25].

## RESULTS

### Sociodemographic and clinical characteristics of people with epilepsy

1343 people with epilepsy were included. 12.7% reported suicidal ideation. 6.1% had a hospital admission following suicide attempt (Table 1). There was a similar proportion of males and females, and the predominant ethnicity was White. 57% had at least one neuropsychiatric comorbidity, the most common being Pervasive Developmental Disorders or Intellectual Disability (31.9%). Other common comorbidities were Anxiety or stress-related disorders (12%), and Depression or Bipolar Disorders (12%).

**Table 1.**
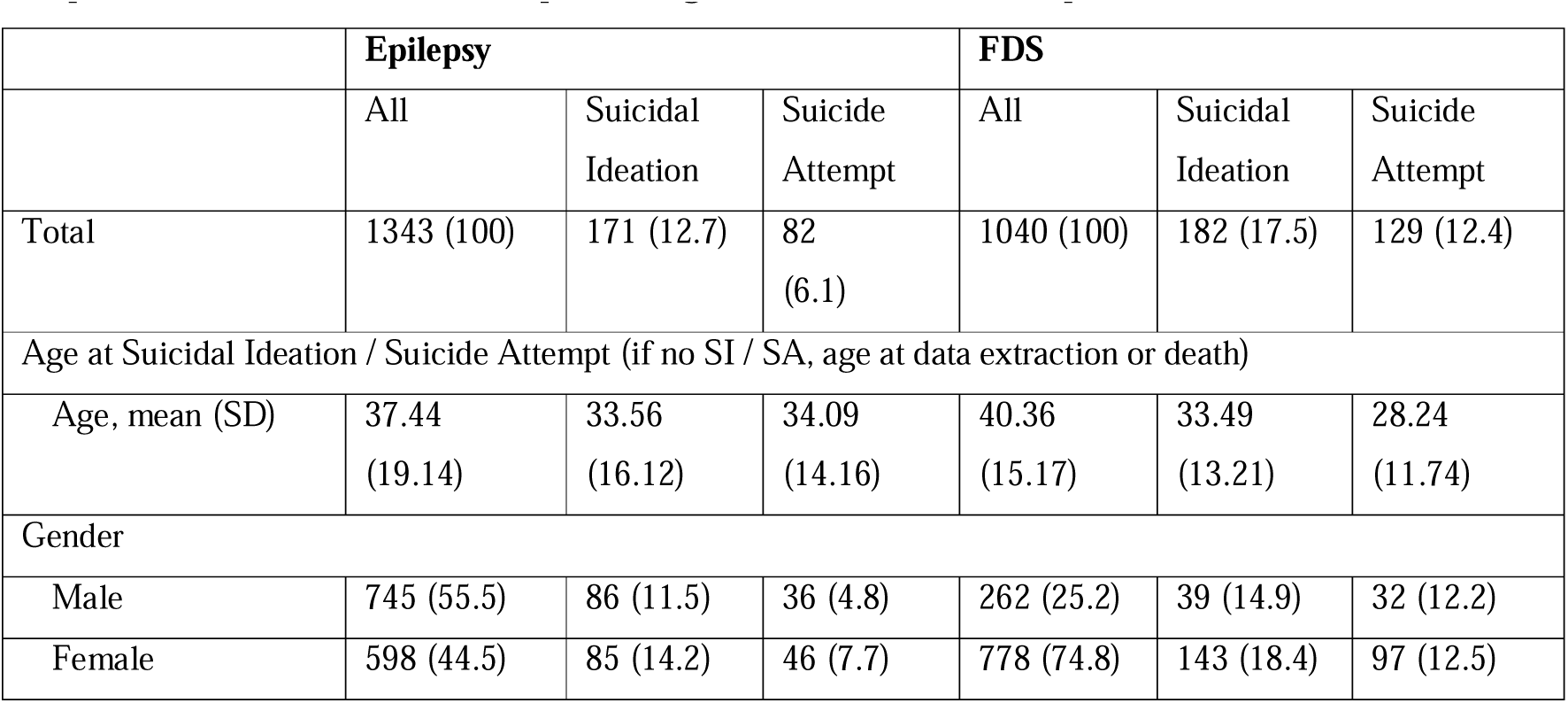
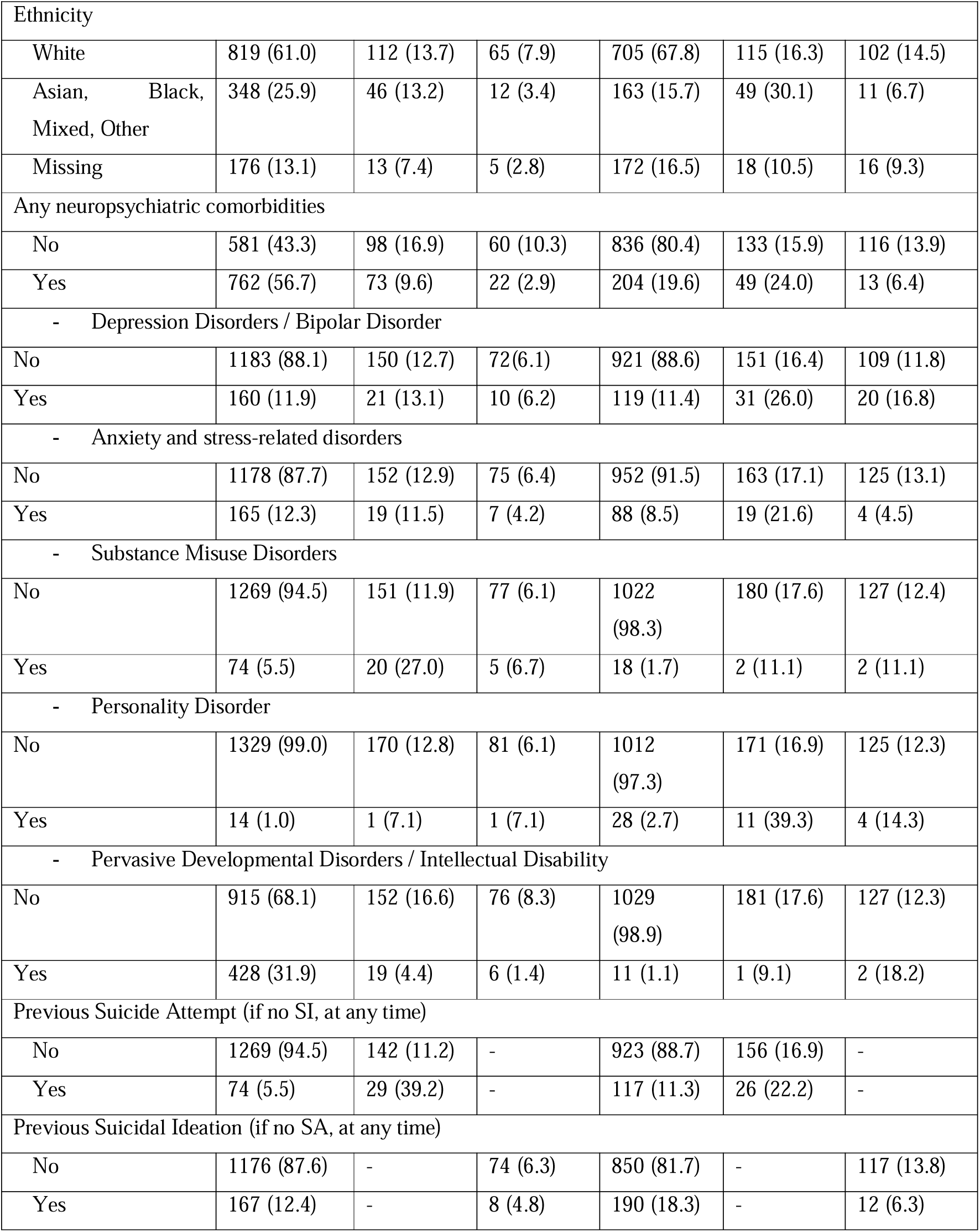
Sociodemographic and clinical characteristics of people with epilepsy and people with FDS seen at South London and Maudsley Hospital between 01/01/2007 and 18/06/2021. Reported are number of cases (percentages) unless otherwise specified.

|  | Epilepsy |  |  | FDS |  |  |
| --- | --- | --- | --- | --- | --- | --- |
|  | All | Suicidal Ideation | Suicide Attempt | All | Suicidal Ideation | Suicide Attempt |
| Total | 1343 (100) | 171 (12.7) | 82 (6.1) | 1040 (100) | 182 (17.5) | 129 (12.4) |
| Age at Suicidal Ideation / Suicide Attempt (if no SI / SA, age at data extraction or death) |  |  |  |  |  |  |
| Age, mean (SD) | 37.44 (19.14) | 33.56 (16.12) | 34.09 (14.16) | 40.36 (15.17) | 33.49 (13.21) | 28.24 (11.74) |
| Gender |  |  |  |  |  |  |
| Male | 745 (55.5) | 86 (11.5) | 36 (4.8) | 262 (25.2) | 39 (14.9) | 32 (12.2) |
| Female | 598 (44.5) | 85 (14.2) | 46 (7.7) | 778 (74.8) | 143 (18.4) | 97 (12.5) |
| Ethnicity |  |  |  |  |  |  |
| White | 819 (61.0) | 112 (13.7) | 65 (7.9) | 705 (67.8) | 115 (16.3) | 102 (14.5) |
| Asian, Black, Mixed, Other | 348 (25.9) | 46 (13.2) | 12 (3.4) | 163 (15.7) | 49 (30.1) | 11 (6.7) |
| Missing | 176 (13.1) | 13 (7.4) | 5 (2.8) | 172 (16.5) | 18 (10.5) | 16 (9.3) |
| Any neuropsychiatric comorbidities |  |  |  |  |  |  |
| No | 581 (43.3) | 98 (16.9) | 60 (10.3) | 836 (80.4) | 133 (15.9) | 116 (13.9) |
| Yes | 762 (56.7) | 73 (9.6) | 22 (2.9) | 204 (19.6) | 49 (24.0) | 13 (6.4) |
| - Depression Disorders / Bipolar Disorder |  |  |  |  |  |  |
| No | 1183 (88.1) | 150 (12.7) | 72 (6.1) | 921 (88.6) | 151 (16.4) | 109 (11.8) |
| Yes | 160 (11.9) | 21 (13.1) | 10 (6.2) | 119 (11.4) | 31 (26.0) | 20 (16.8) |
| - Anxiety and stress-related disorders |  |  |  |  |  |  |
| No | 1178 (87.7) | 152 (12.9) | 75 (6.4) | 952 (91.5) | 163 (17.1) | 125 (13.1) |
| Yes | 165 (12.3) | 19 (11.5) | 7 (4.2) | 88 (8.5) | 19 (21.6) | 4 (4.5) |
| - Substance Misuse Disorders |  |  |  |  |  |  |
| No | 1269 (94.5) | 151 (11.9) | 77 (6.1) | 1022 (98.3) | 180 (17.6) | 127 (12.4) |
| Yes | 74 (5.5) | 20 (27.0) | 5 (6.7) | 18 (1.7) | 2 (11.1) | 2 (11.1) |
| - Personality Disorder |  |  |  |  |  |  |
| No | 1329 (99.0) | 170 (12.8) | 81 (6.1) | 1012 (97.3) | 171 (16.9) | 125 (12.3) |
| Yes | 14 (1.0) | 1 (7.1) | 1 (7.1) | 28 (2.7) | 11 (39.3) | 4 (14.3) |
| - Pervasive Developmental Disorders / Intellectual Disability |  |  |  |  |  |  |
| No | 915 (68.1) | 152 (16.6) | 76 (8.3) | 1029 (98.9) | 181 (17.6) | 127 (12.3) |
| Yes | 428 (31.9) | 19 (4.4) | 6 (1.4) | 11 (1.1) | 1 (9.1) | 2 (18.2) |
| Previous Suicide Attempt (if no SI, at any time) |  |  |  |  |  |  |
| No | 1269 (94.5) | 142 (11.2) | - | 923 (88.7) | 156 (16.9) | - |
| Yes | 74 (5.5) | 29 (39.2) | - | 117 (11.3) | 26 (22.2) | - |
| Previous Suicidal Ideation (if no SA, at any time) |  |  |  |  |  |  |
| No | 1176 (87.6) | - | 74 (6.3) | 850 (81.7) | - | 117 (13.8) |
| Yes | 167 (12.4) | - | 8 (4.8) | 190 (18.3) | - | 12 (6.3) |

### Sociodemographic and clinical characteristics of people with FDS

1040 people with FDS were included. 17.5% reported suicidal ideation. 12.4% had a hospital admission following suicide attempt (Table 1). 75% were females, and the predominant ethnicity was White. A neuropsychiatric comorbidity was present in approximately 20%, the most common being Depression or Bipolar Disorder (11%) and Anxiety and stress-related disorders (8.5%).

### Risk and protective factors in people with epilepsy

Results of the bias-reduced Logistic Regression indicated that in people with epilepsy, there is a significant non-linear association between age and suicidal ideation; odds were stable until the knot location, i.e., age 33 (OR: 1.03; CI: 1.00 – 1.05) before declining (OR: 0.94; CI: 0.90 – 0.98; Table 2; Figure 1). Age had a significant quadratic association with suicide-attempt related admissions, with risk increasing until age 33/knot (OR: 1.06; CI: 1.02 – 1.11) before declining (OR: 0.88; CI: 0.82 – 0.94; Table 3; Figure 1). Whilst both genders had similar odds of experiencing suicidal ideation (OR: 1.27; CI: 0.92 – 1.75; Table 2), the odds of females having a first admission for suicide attempt were significantly increased as compared to males (OR: 1.64; CI: 1.05 – 2.56; Table 3). Whilst ethnicity was not significantly associated with suicidal ideation (OR:0.96; CI: 0.66 – 1.38; Table 2), the odds of people of White ethnicity being admitted following suicide attempt were significantly elevated (1/0.45 = 2.22 times) as compared to other ethnic backgrounds (Asian, Black, Mixed, Multiple and Other; OR: 0.45; CI: 0.24 – 0.85; Table 3). The odds of people with a Substance Misuse Disorder of experiencing suicidal ideation were 2.67 times the odds of people without (OR: 2.67; CI: 1.61 – 4.74; Table 2). The presence of a Pervasive Developmental Disorders or Intellectual Disability was associated with a significantly reduced likelihood of both suicidal ideation (OR: 0.24; CI: 0.15 – 0.39; Table 2) and suicide attempt-related admission (OR: 0.16; CI: 0.07 – 0.35; Table 3). There was no association between our outcomes and a diagnosis of Depression or Bipolar Disorder, or Anxiety or stress-related disorders (Table 2, Table 3). Reporting suicidal ideation did not affect the likelihood of subsequent suicide attempt-related admission (OR: 1.04; CI: 0.49 – 2.18; Table 3). Results for the D3 likelihood ratio test are in Supplementary Material. R^2^ values were low; predictor variables explained between 0% and 3.8% of the variance in the outcomes studied (Supplementary Tables 2-3).

**Figure 1.**
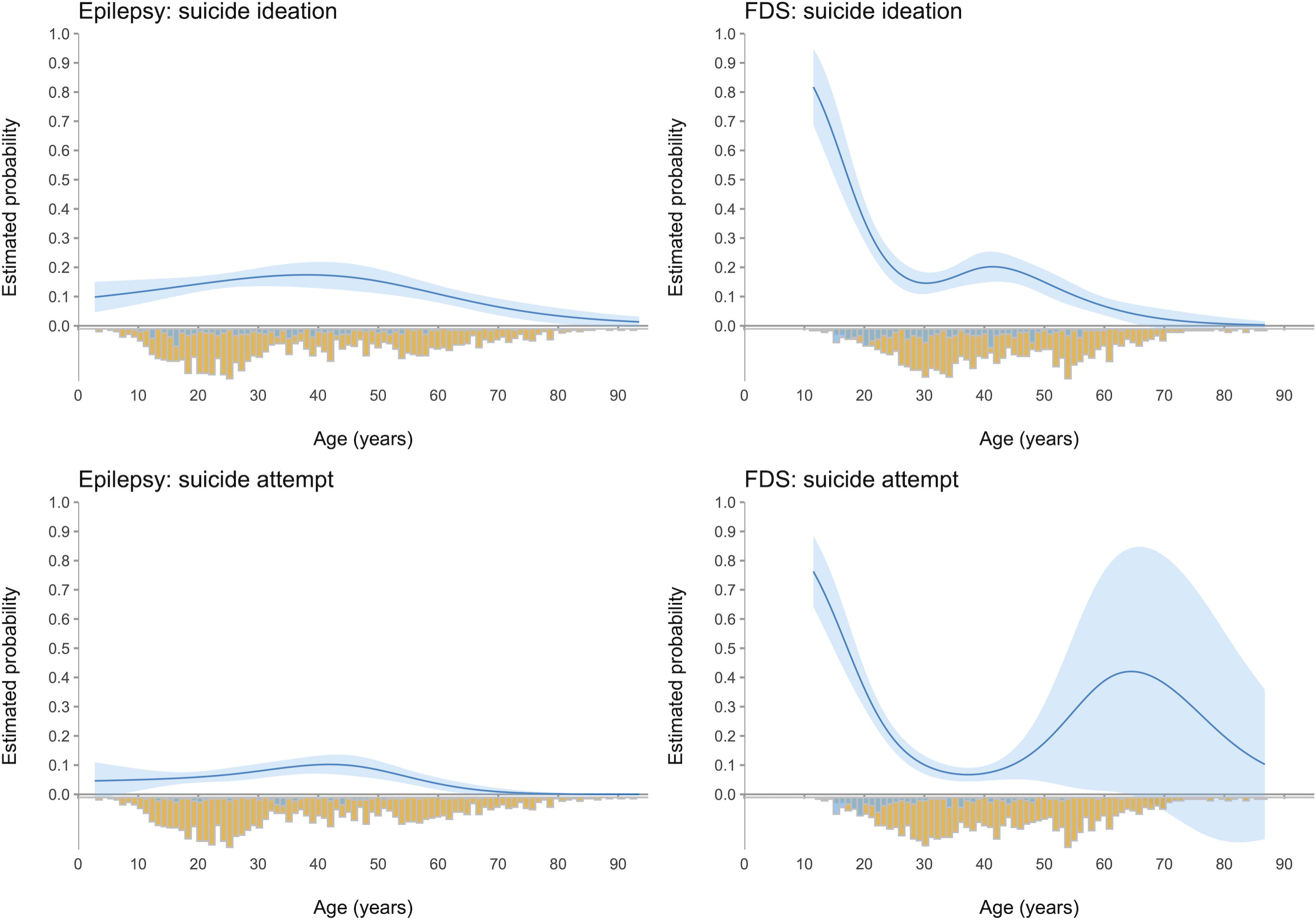
Loess smoothed (local non-linear regression) curves showing probability of suicidal ideation and suicide attempt-related hospitalisation by age per each diagnostic group. (top left) Probability of suicidal ideation by age in epilepsy; (bottom left) Probability of suicide attempt-related admission by age in epilepsy; (top right) Probability of suicidal ideation by age in FDS; (bottom right) Probability of suicide attempt-related admission by age in FDS. On the negative side of the y axis are depicted the age distributions (number of cases) stratified by presence of outcome (blue: suicidal ideation/suicide attempt present; yellow: suicidal ideation/suicide attempt absent).

**Table 2.** Results from bias-reduced Logistic Regression analysis on **suicidal ideation in people with epilepsy,** presented as Beta values and associated Standard Errors (SE), Odds Ratios (ORs) with 95% Confidence Intervals (CIs), and significance value (p).

|  | B (SE) | Odds Ratio | 95% CI for<br>OR (lower) | 95% CI for<br>OR (upper) | p |
| --- | --- | --- | --- | --- | --- |
| Age at SI (linear)<sup>1</sup> | -0.01 (0.0) | 0.99 | 0.98 | 1.00 | 0.008 |
| Age at SI (non-linear 1, age < 32.8)<sup>1</sup> | 0.02 (0.01) | 1.03 | 1.00 | 1.05 | 0.068 |
| Age at SI (non-linear 2, age > 32.8)<sup>1</sup> | -0.06 (0.02) | 0.94 | 0.9 | 0.98 | 0.004 |
| Gender<sup>2</sup> | 0.24 (0.16) | 1.27 | 0.92 | 1.75 | 0.145 |
| Ethnicity<sup>3</sup> | -0.04 (0.19) | 0.96 | 0.66 | 1.38 | 0.815 |
| Any neuropsychiatric comorbidities<sup>4</sup> | -0.64 (0.17) | 0.53 | 0.38 | 0.73 | <0.001 |
| - Depression / Bipolar <sup>4a</sup> | 0.06 (0.25) | 1.06 | 0.64 | 1.73 | 0.83 |
| - Anxiety / stress-related <sup>4b</sup> | -0.12 (0.26) | 0.89 | 0.53 | 1.47 | 0.64 |
| - Substance Misuse <sup>4c</sup> | 1.02 (0.28) | 2.67 | 1.61 | 4.74 | <0.001 |
| - PDD / ID <sup>4d</sup> | -1.43 (0.25) | 0.24 | 0.15 | 0.39 | <0.001 |

<sup>1</sup> Model just including age as predictor.
<sup>2</sup> Model just includes gender as a predictor, with male as reference category.
<sup>3</sup> Model just includes ethnicity as a predictor, with white as reference category.
<sup>4</sup> Model includes ethnicity and gender as covariates.
<sup>4a</sup> Model including ethnicity, gender, anxiety, DD/ID as covariates.
<sup>4b</sup> Model including ethnicity, gender, Depressive Disorders, Substance Misuse as covariates.
<sup>4c</sup> Model including ethnicity and Anxiety Disorders as covariates.
<sup>4d</sup> Model only including PDD/ID as predictor.

**Table 3.**
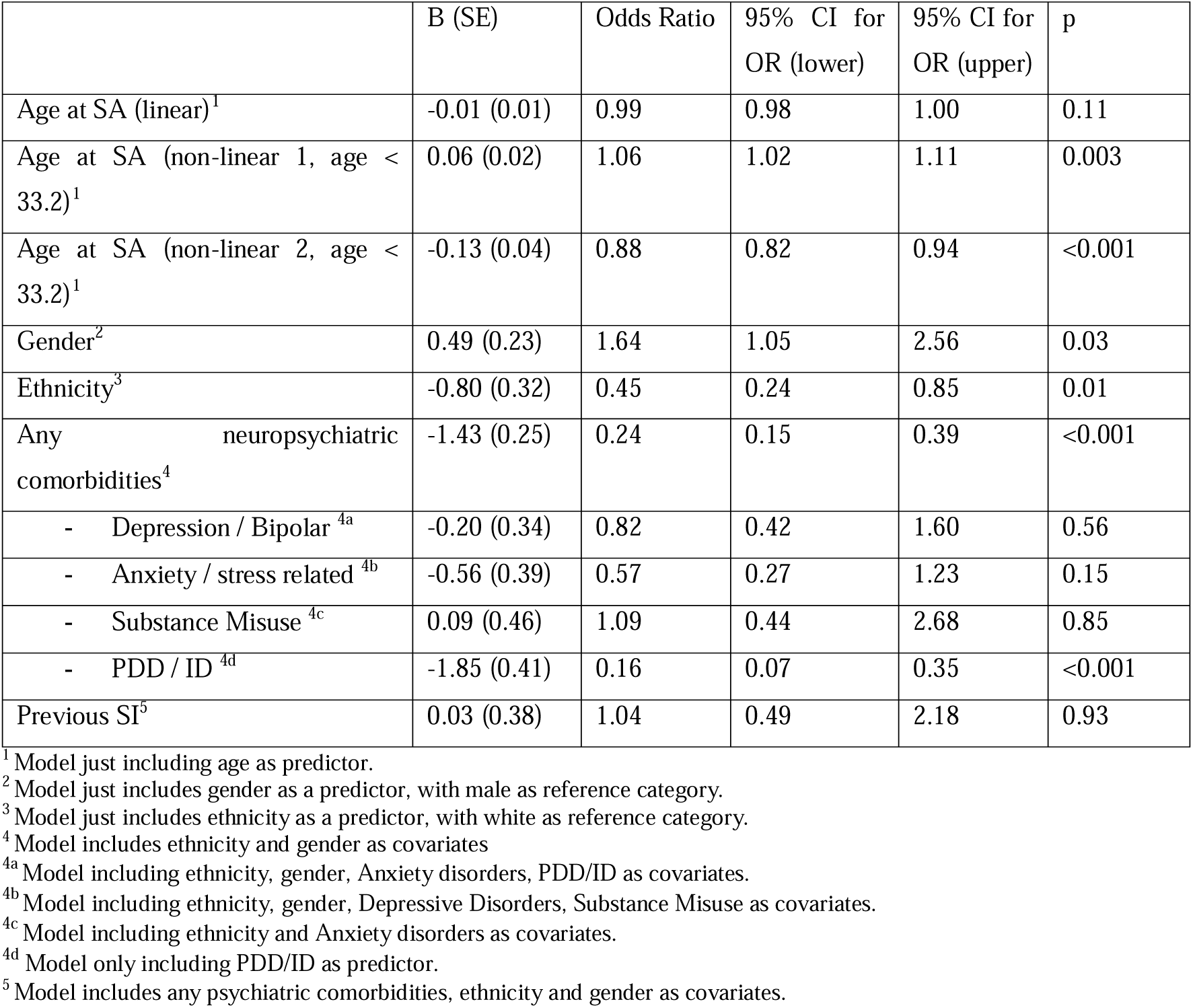
Results from bias-reduced Logistic Regression analysis on **suicide attempt-related admissions in people with epilepsy,** presented as Beta values and associated Standard Errors (SE), Odds Ratios (ORs) with 95% Confidence Intervals (CIs), and significance value (p).

|  | B (SE) | Odds Ratio | 95% CI for OR (lower) | 95% CI for OR (upper) | p |
| --- | --- | --- | --- | --- | --- |
| Age at SA (linear)<sup>1</sup> | -0.01 (0.01) | 0.99 | 0.98 | 1.00 | 0.11 |
| Age at SA (non-linear 1, age < 33.2)<sup>1</sup> | 0.06 (0.02) | 1.06 | 1.02 | 1.11 | 0.003 |
| Age at SA (non-linear 2, age < 33.2)<sup>1</sup> | -0.13 (0.04) | 0.88 | 0.82 | 0.94 | <0.001 |
| Gender<sup>2</sup> | 0.49 (0.23) | 1.64 | 1.05 | 2.56 | 0.03 |
| Ethnicity<sup>3</sup> | -0.80 (0.32) | 0.45 | 0.24 | 0.85 | 0.01 |
| Any neuropsychiatric comorbidities<sup>4</sup> | -1.43 (0.25) | 0.24 | 0.15 | 0.39 | <0.001 |
| - Depression / Bipolar <sup>4a</sup> | -0.20 (0.34) | 0.82 | 0.42 | 1.60 | 0.56 |
| - Anxiety / stress related <sup>4b</sup> | -0.56 (0.39) | 0.57 | 0.27 | 1.23 | 0.15 |
| - Substance Misuse <sup>4c</sup> | 0.09 (0.46) | 1.09 | 0.44 | 2.68 | 0.85 |
| - PDD / ID <sup>4d</sup> | -1.85 (0.41) | 0.16 | 0.07 | 0.35 | <0.001 |
| Previous SI<sup>5</sup> | 0.03 (0.38) | 1.04 | 0.49 | 2.18 | 0.93 |

<sup>1</sup> Model just including age as predictor.
<sup>2</sup> Model just includes gender as a predictor, with male as reference category.
<sup>3</sup> Model just includes ethnicity as a predictor, with white as reference category.
<sup>4</sup> Model includes ethnicity and gender as covariates
<sup>4a</sup> Model including ethnicity, gender, Anxiety disorders, PDD/ID as covariates.
<sup>4b</sup> Model including ethnicity, gender, Depressive Disorders, Substance Misuse as covariates.
<sup>4c</sup> Model including ethnicity and Anxiety disorders as covariates.
<sup>4d</sup> Model only including PDD/ID as predictor.
<sup>5</sup> Model includes any psychiatric comorbidities, ethnicity and gender as covariates.

### Risk and protective factors in people with FDS

Results of the bias-reduced Logistic Regression indicated that as age of people with FDS increases, there is a significant decrease in the odds of suicidal ideation (age 33/knot location: OR: 0.85; CI: 0.81 – 0.90; Table 4; Figure 1) and suicide-attempt related hospitalisation (age 32/knot location: OR: 0.81; CI: 0.77 – 0.86; Table 5; Figure 1). After age 33 or 32, risk increases again until age 47 (for ideation: OR:1.73; CI: 1.41 – 2.14; for attempt: OR: 1.91; CI: 1.45 – 2.52) before declining (for ideation: OR: 0.20; CI: 0.11 – 0.38; for attempt: OR: 0.16; CI: 0.07 – 0.38). Both genders had similar odds for suicidality (Table 4; Table 5). The odds of people of Asian, Black, Mixed, Multiple and Other ethnicities of experiencing suicidal ideation were significantly higher (2.4 times) than those of people of White ethnicity (OR: 2.40; CI: 1.63 – 3.53; Table 4). However, people of White ethnicity were 1/0.49 = 2.0 times more likely to have an admission following a suicide attempt (OR: 0.49; CI: 0.26 – 0.92; Table 5). The odds of people with comorbid Depression or Bipolar Disorder of experiencing suicidal ideation were significantly higher compared to those without these diagnoses (OR: 1.91; CI: 1.22 – 3.00; Table 4). Odds were significantly higher also for people with comorbid Personality Disorder (OR: 3.01; CI: 1.37 – 6.63; Table 4). There was no association between suicidal ideation and a diagnosis of Anxiety or stress-related disorders (Table 4). In relation to suicide attempt-related admissions, it was only possible to explore the association with a diagnosis of Depression or Bipolar Disorder (non-significant association; Table 5) due to low cell counts for the other comorbidities. Reporting previous suicidal ideation was not associated to subsequent suicide attempt-related admission (OR: 0.62; CI: 0.33 – 1.16; Table 5). Predictor variables explained between 0% and 15.7% of the variance in the outcomes studied (Supplementary Tables 4-5).

**Table 4.** Results from bias-reduced Logistic Regression analysis on **suicidal ideation in people with FDS,** presented as Beta values and associated Standard Errors (SE), Odds Ratios (ORs) with 95% Confidence Intervals (CIs), and significance value (p).

|  | B (SE) | Odds Ratio | 95% CI for<br>OR (lower) | 95% CI for<br>OR (upper) | p |
| --- | --- | --- | --- | --- | --- |
| Age at SI (linear)<sup>1</sup> | -0.05 (0.01) | 0.96 | 0.94 | 0.97 | <0.001 |
| Age at SI (non-linear 1; age < 33)<sup>1</sup> | -0.16 (0.03) | 0.85 | 0.81 | 0.9 | <0.001 |
| Age at SI (non-linear 2; 33 < age < 47)<sup>1</sup> | 0.55 (0.11) | 1.73 | 1.41 | 2.14 | <0.001 |
| Age at SI (non-linear 3; age > 47)<sup>1</sup> | -1.59 (0.32) | 0.20 | 0.11 | 0.38 | <0.001 |
| Gender<sup>2</sup> | 0.25 (0.20) | 1.28 | 0.87 | 1.88 | 0.21 |
| Ethnicity<sup>3</sup> | 0.87 (0.20) | 2.40 | 1.63 | 3.53 | <0.001 |
| Any neuropsychiatric comorbidities<sup>4</sup> | 0.52 (0.19) | 1.68 | 1.15 | 2.45 | 0.007 |
| - Depression / Bipolar <sup>4a</sup> | 0.65 (0.23) | 1.91 | 1.22 | 3.00 | 0.005 |
| - Anxiety / stress-related <sup>4b</sup> | 0.29 (0.28) | 1.34 | 0.78 | 2.29 | 0.29 |
| - Personality Disorder <sup>4c</sup> | 1.10 (0.40) | 3.01 | 1.37 | 6.63 | 0.006 |

<sup>1</sup> Model just including age as predictor.
<sup>2</sup> Model just includes gender as a predictor, with male as reference category.
<sup>3</sup> Model just includes ethnicity as a predictor, with white as reference category.
<sup>4</sup> Model includes ethnicity and gender as covariates.
<sup>4a</sup> Model including ethnicity, gender, Anxiety disorders, PDD/ID as covariates.
<sup>4b</sup> Model including ethnicity, gender, Depressive Disorders, Substance Misuse as covariates.
<sup>4c</sup> Model including ethnicity and gender as covariates.

**Table 5.** Results from bias-reduced Logistic Regression analysis on **suicide attempt-related admissions in people with FDS,** presented as Beta values and associated Standard Errors (SE), Odds Ratios (ORs) with 95% Confidence Intervals (CIs), and significance value (p).

|  | B (SE) | Odds Ratio | 95% CI for OR (lower) | 95% CI for OR (upper) | p |
| --- | --- | --- | --- | --- | --- |
| Age at SA (linear)<sup>1</sup> | -0.08 (0.01) | 0.92 | 0.90 | 0.94 | <0.001 |
| Age at SA (non-linear 1; age < 32)<sup>1</sup> | -0.21 (0.03) | 0.81 | 0.77 | 0.86 | <0.001 |
| Age at SA (non-linear 2; 32 < age < 47)<sup>1</sup> | 0.65 (0.14) | 1.91 | 1.45 | 2.52 | <0.001 |
| Age at SA (non-linear 3; age > 47)<sup>1</sup> | -1.81 (0.43) | 0.16 | 0.07 | 0.38 | <0.001 |
| Gender<sup>2</sup> | 0.01 (0.22) | 1.01 | 0.66 | 1.55 | 0.95 |
| Ethnicity<sup>3</sup> | -0.72 (0.32) | 0.49 | 0.26 | 0.92 | 0.026 |
| Neuropsychiatric comorbidities |  |  |  |  |  |
| - Depression/Bipolar <sup>4a</sup> | 0.07 (0.26) | 1.07 | 0.64 | 1.78 | 0.79 |
| Previous SI | -0.48 (0.32) | 0.62 | 0.33 | 1.16 | 0.13 |

<sup>1</sup> Model just including age as predictor.
<sup>2</sup> Model just includes gender as a predictor, with male as reference category.
<sup>3</sup> Model just includes ethnicity as a predictor, with white as reference category.
<sup>4a</sup> Model including ethnicity, gender, Anxiety Disorders, PDD/ID as covariates.
<sup>5</sup> Model includes any psychiatric comorbidities, ethnicity and gender as covariates.

## DISCUSSION

This work sought to identify the sociodemographic and comorbidity correlates of suicidal ideation and suicide attempt-related hospital admissions in large cohorts of people with FDS or epilepsy. We studied group-specific associations with age, gender, ethnicity, and comorbid neuropsychiatric diagnoses. We present novel findings for risk factors in the FDS population which have not been previously studied.

We identify factors that have similar relationships with suicidality across the two groups, indicating that these are more likely to be general risk factors, rather than specific to a particular clinical population.

For both cohorts, ethnicity accounted for a small proportion of the observed variance in suicide attempt-related hospitalisation (0.5-0.6%). However, a consistent association was observed; people of Asian, Black, Mixed, Multiple or Other backgrounds had significantly reduced odds for such hospitalisations (51-55% reduction) compared to people of White ethnicity. These results align with the largest UK community study[26], highlighting that people of ethnic minorities are half as likely to receive medical attention following suicide attempt than people of white ethnicity. In our study, this was observed despite ethnic minorities having significantly higher odds of reporting suicidal ideation in part of our sample (FDS group, OR: 2.4). It has been suggested that scarcer service accessibility or fear of stigma may deter individuals from ethnic minorities seeking medical attention after suicide attempts[26]. Further research is required to understand this; if appropriate, efforts should be implemented to reduce stigmatisation and improve service access for ethnic minorities following suicide attempts.

In both cohorts, reporting suicidal ideation was not associated with a higher likelihood of subsequent suicide attempt-related admission. Our measure of suicidal ideation relied on Natural Language Processing of clinical notes rather than systematic assessment, and it is uncertain if ideation was reported elsewhere. Nevertheless, our results highlight the difficulty of predicting suicide attempts amongst people experiencing suicidal ideation, as most do not progress to actual attempts[27,28].

We have identified a number of risk and protective factors for suicidality that are unique to each diagnostic group.

In people with FDS, both genders had comparable odds of reporting suicidal ideation and of being hospitalised following a suicide attempt. This indicates that although the FDS group has a high prevalence (75%) of females, both genders are at similar suicidality risk. This is a novel finding which has not been previously reported. In people with epilepsy, no gender effect was observed for suicidal ideation. However, females had significantly higher odds of having a suicide attempt-related admission (OR: 1.64; 0.4% variance explained), in line with previous evidence[11,28].

In people with FDS, age was significantly associated with a first instance of suicidal ideation and a first suicide attempt-related hospitalisation, each explaining 8.8% and 15.7% of the outcome variance respectively. Consistent with previous studies in the general population[29], we observed that adolescence and early adulthood are periods of greatest risk for onset of suicidality in FDS, with risk varying non-linearly as a function of age.

In epilepsy, age had a significant non-linear association with both suicidality outcomes, explaining 1.1 – 1.2% of their variance. Consistent with the probability distribution observed in this study, previous evidence indicates that people with epilepsy tend to complete suicide later in life (in their 40s) compared to those without epilepsy[30,31]. This might relate to the highest prevalence of epilepsy occurring in the 35 to 64 age group[31], although our sample distribution does not support this, as it includes a high proportion of cases under 30 years old. Suicidality in epilepsy is multifaceted and may have distinct underlying mechanisms compared to those driving the suicidality peak observed in adolescence and early adulthood in FDS or the general population. Epilepsy-related and psychosocial factors, treatment responsiveness, and the shared neurobiological substrate between depression and epilepsy[32] may contribute to a more chronic vulnerability to suicidality later in life.

Analysis of five comorbidity classes revealed that suicidality in FDS and epilepsy is associated with a distinct pattern of neuropsychiatric comorbidities.

In people with epilepsy, those with a comorbid diagnosis of Substance Misuse had 167% higher odds of reporting suicidal ideation (1.1% variance explained), consistently with reports from a recent meta-analysis[33]. As previously reported[12], we found that a strong protective factor against suicidal ideation (76% reduction in odds) and suicide-attempt related admissions (84% reduction) was a diagnosis of moderate to profound Intellectual Disability or Pervasive Developmental Disorder such as Autism, Rett syndrome, and Asperger syndrome. This explained 2-2.9% of the variance in suicidality outcomes. An impaired communication of thoughts and emotions might contribute to underreported suicidal ideation in these populations; these may also have only a partial understanding of the concepts of life and death, which serves as the basis of forming suicide intent[34]. The reasons driving this finding should be matter of further study.

In people with epilepsy, diagnoses of Depression, Bipolar Disorder, Anxiety, or stress-related disorders (including OCD and PTSD) were not associated with suicidality. Whilst meta-analysis results suggest an overall weak association between anxiety, related disorders and suicidality[35], our finding of a lack of association for Depressive Disorders is surprising, as these have been reported as the major risk factor for suicidal ideation in epilepsy[10]. However, recent meta-analysis results also suggest that depression might not effectively predict suicide attempts[27]. Our results may be due to sample-specific characteristics, to sampling from a large tertiary mental health hospital or to limitations in capturing relevant factors such as disorder severity or hopelessness using ICD-10 codes for depression[36]. These results should be better understood in future meta-analyses.

In people with FDS, the odds of experiencing suicidal ideation were 200% higher in those with a comorbid diagnosis of Personality Disorder, and this accounted for 2.9% of the variance. Suicidality is a central feature of the Personality Disorders subtypes strongly associated with FDS[37], such as Borderline Personality Disorder. The risk of reporting suicidal ideation was also higher in the presence of a diagnosis of Depression or Bipolar Disorder (91% increase in odds, 3% variance explained), although this did not translate to an increased risk of suicide attempt-related hospitalisation in this group. Again, as Depressive Disorders are strongly associated with suicidality, these results do not imply an absence of increased risk and should be considered in future aggregative studies for a more comprehensive interpretation. It was not possible to estimate the association between suicide attempt-related admissions and a comorbid diagnosis of Anxiety or stress-related disorders, Personality Disorders, Substance Misuse Disorder, or Pervasive Developmental Disorder due to low numbers in this group.

Figure 2 includes a summary of clinically relevant study findings for the FDS group.

**Figure 2.**
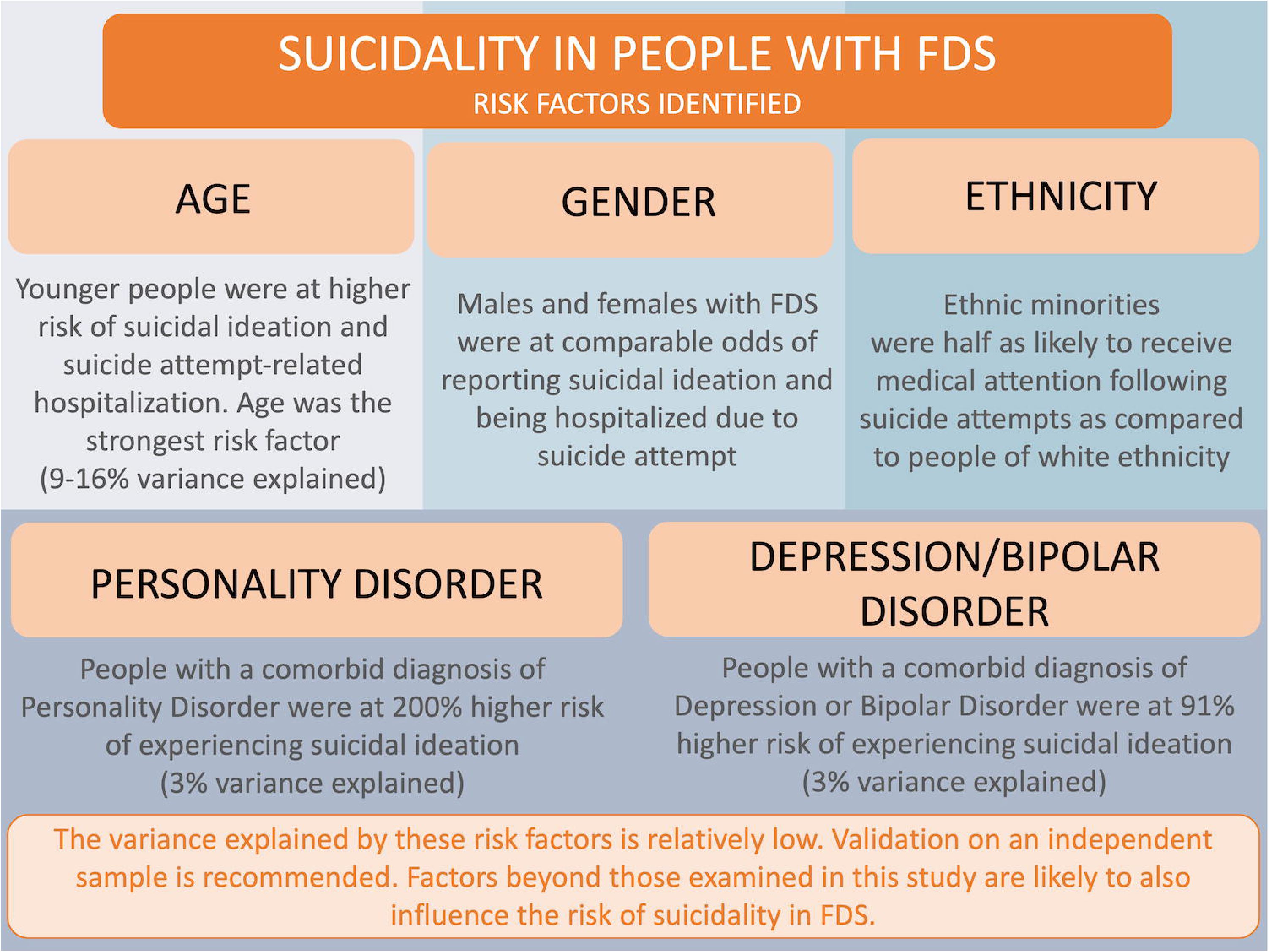
Summary of main findings on risk factors for suicidality in people with FDS.

This study’s major strength is the sizable patient sample (n = 2383) from the UK largest tertiary mental health hospital, spanning 15 years. Linkage to national admission data from the HES ensured that hospitalisations could be captured independently from location of the admitting hospital. However, non-admissions following suicide attempt, estimated to be 50% of those attempting suicide[38], were not considered. Cases of suicidal ideation were not captured if unreported or undocumented during hospital appointments at our site. Our NLP app for identifying ideation had 60 - 71% recall and an estimated 91.7% precision; whilst all false positives were corrected prior to analyses, a proportion of relevant instances might have been missed. Severity and frequency of ideation was not assessed.

Due to the study’s retrospective nature, information on how the diagnoses of epilepsy or FDS were arrived at was not available, and a degree of misdiagnosis might be present; for epilepsy, this is estimated to be between 4.6 and 5.6% following specialist review[39]. We expect minimal misdiagnosis rates for FDS due to the hospital’s high specialisation for FDS. Our use of ICD-10 codes for identifying comorbid disorders might underestimate their true prevalence.

Our patient cohort might differ from that of community studies because of sampling from a tertiary mental health hospital. For example, our epilepsy sample showed a higher prevalence of at least one of the psychiatric comorbidities under study (57%), compared to the typically reported 30% prevalence for psychiatric comorbidities in epilepsy[40]. Results may also vary under different theoretical assumptions (diagrams).

## CONCLUSION

This is the first study to systematically examine the correlates of suicidality in people with FDS. We also examined a large sample of people with epilepsy. Collectively, results suggest that shared sociodemographic and clinical risk factors, as well as disorder-specific risk factors for suicidality, can be identified. Characteristics such as ethnicity had similar, systematic associations with suicidality in both groups. A pattern of association that was unique to each clinical population was found for gender, age, and comorbidity profile.

Whilst correlates of suicidality such as disorder-specific comorbidity profiles will be useful to identify groups at higher risk in clinical settings, factors such as ethnicity are suitable targets for population-based health strategies and preventive programs.

## Supporting information

RECORD Checklist

## ACKNOWLEDGEMENTS

We would like to acknowledge Dr Elena Bernardini for a discussion on the correlates and theories of suicidality.

We would like to acknowledge the Bergqvist Charitable Trust and the Psychiatry Research Trust for their financial support.

This study represents independent research partly funded by the National Institute for Health and Care Research (NIHR) Biomedical Research Centre at South London and Maudsley NHS Foundation Trust and King’s College London. The views expressed are those of the author(s) and not necessarily those of the NHS, the NIHR or the Department of Health and Social Care. We would like to acknowledge the Office for National Statistics (ONS) as the provider of the Mortality data; those who carried out the original collection and analysis of the data bear no responsibility for their further analysis or interpretation.

Professor Young’s independent research is funded by the National Institute for Health Research (NIHR) Biomedical Research Centre at South London and Maudsley NHS Foundation Trust and King’s College London.

For the purposes of open access, the author has applied a Creative Commons Attribution (CC BY) licence to any Accepted Author Manuscript version arising from this submission.

## AUTHOR CONTRIBUTORSHIP

Concept and design: IF, JH, AHY, PS. Analysis planning: IF, JH. Data collection: IF, IJ. Statistical analyses: JH. Manuscript drafting: IF. Critical manuscript revision: JH, IJ, AHY, PS

## DATA AVAILABILITY STATEMENT

Data may be obtained from a third party and are not publicly available. Access to Clinical Record Interactive Search (CRIS) data used for this study is regulated by the CRIS Oversight Committee and the ‘Oxfordshire C’ Research Ethics approval for secondary analysis of CRIS data (23/SC/0257). Data used for this research cannot be shared without prior approval from the CRIS Oversight Committee. Those interested should contact Robert Stewart (robert.), CRIS academic lead.

## SUPPLEMENTARY MATERIAL

**Supplementary File 1.** Supplementary Figures and Tables

**Supplementary File 2**. STROBE and RECORD checklist

