## Supplementary Material for "Sociodemographic and clinical risk factors for suicidal ideation and suicide attempt in functional/dissociative seizures and epilepsy: a large cohort study"

### **SUPPLEMENTARY FIGURES AND TABLES**

#### Supplementary Figures

Diagrams depicted in Supplementary Figures 1 and 2 were used when estimating the association between our study outcomes and demographic and clinical variables of interest. Diagrams depicted in Supplementary Figures 3 and 4 were used when estimating the association between our study outcomes and each neuropsychiatric comorbidity specifically; this allowed us to further determine the specific drivers of the association between having any neuropsychiatric comorbidity and the study outcomes.

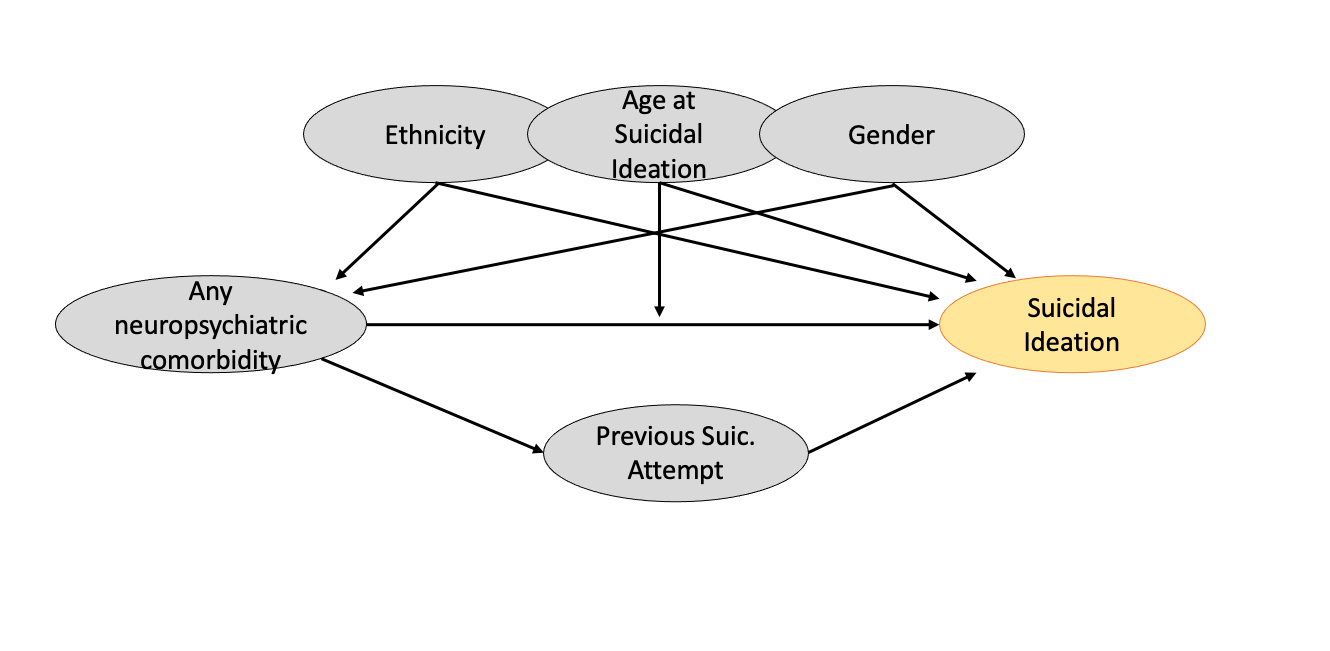

**Supplementary figure 1.** Causal diagram depicting demographic and clinical variables in relation to Suicidal Ideation. An arrow’s direction represents the theoretical direction of causal effect. An arrow pointing to the middle body of another arrow indicates a theoretical moderation effect. Dark grey = predictor variables. Yellow = predicted variable.

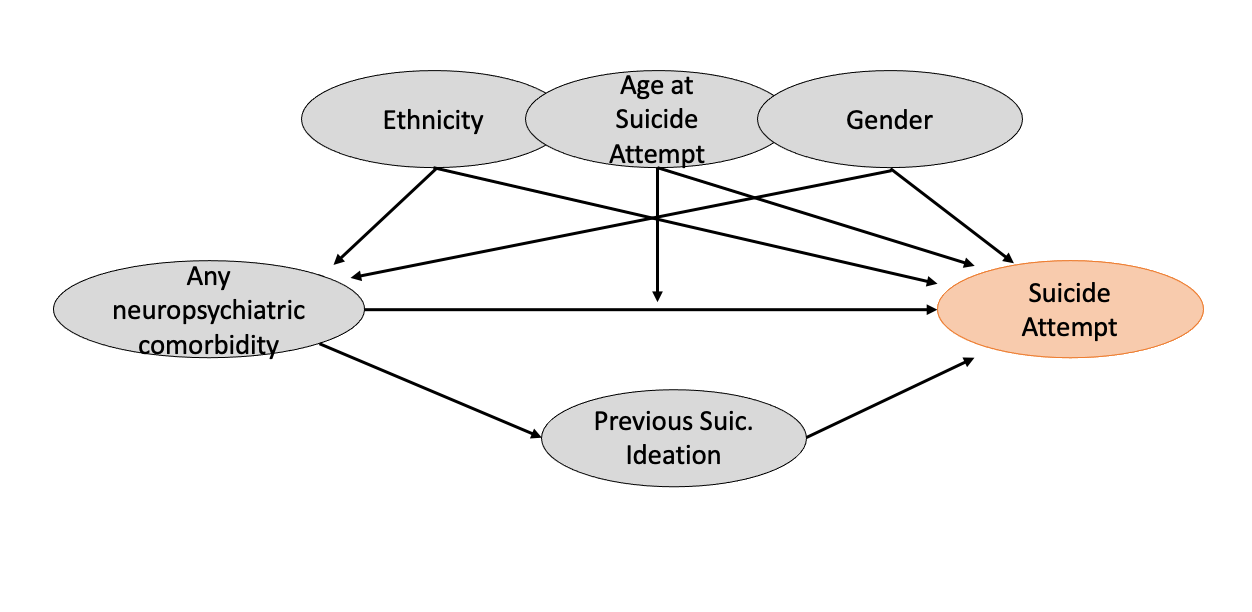

**Supplementary figure 2.** Causal diagram depicting demographic and clinical variables in relation to Suicide Attempt. An arrow’s direction represents the theoretical direction of causal effect. An arrow pointing to the middle body of another arrow indicates a theoretical moderation effect. Dark grey = predictor variables. Red = predicted variable.

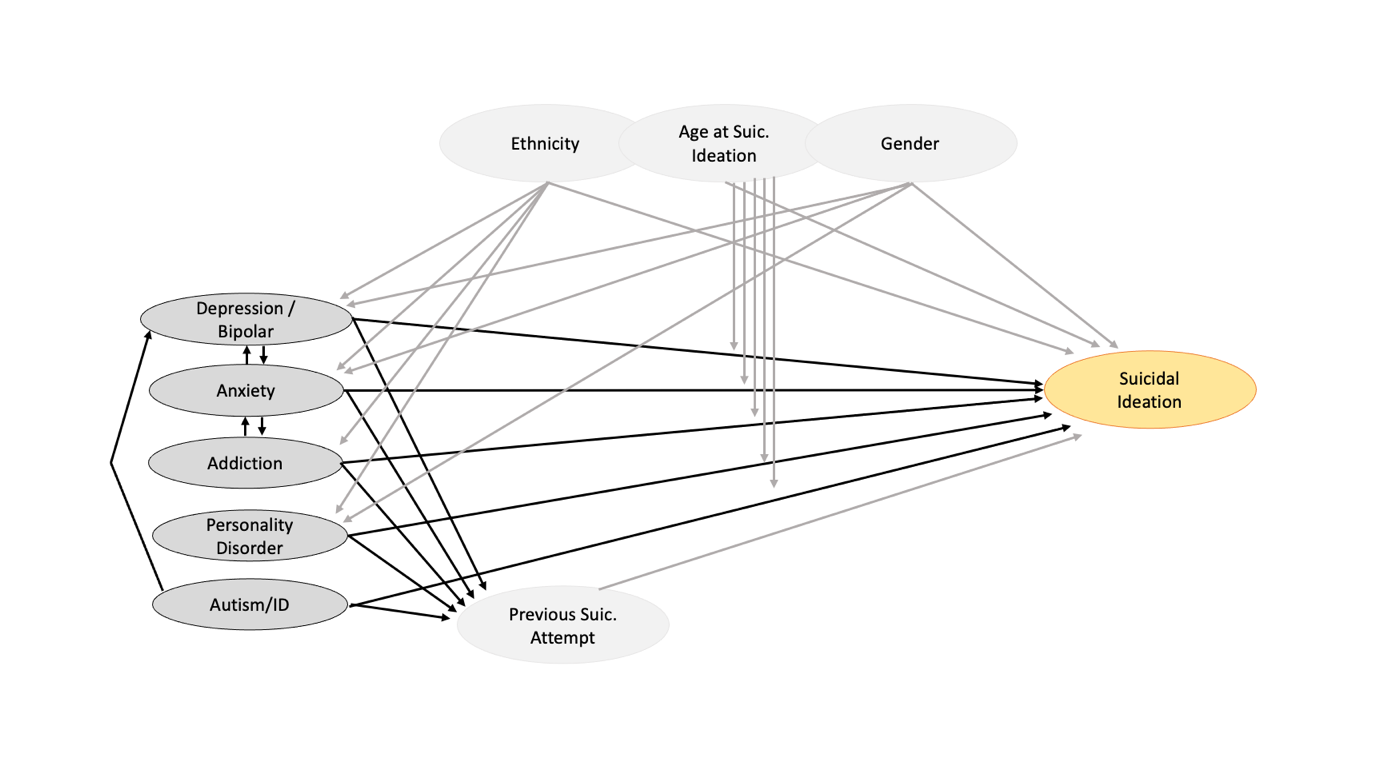

**Supplementary figure 3.** Causal diagram depicting psychiatric comorbidity variables in relation to Suicidal Ideation. An arrow’s direction represents the theoretical direction of causal effect. An arrow pointing to the middle body of another arrow indicates a theoretical moderation effect. Dark grey = predictor variables. Light grey = control variables. Yellow = predicted variable.

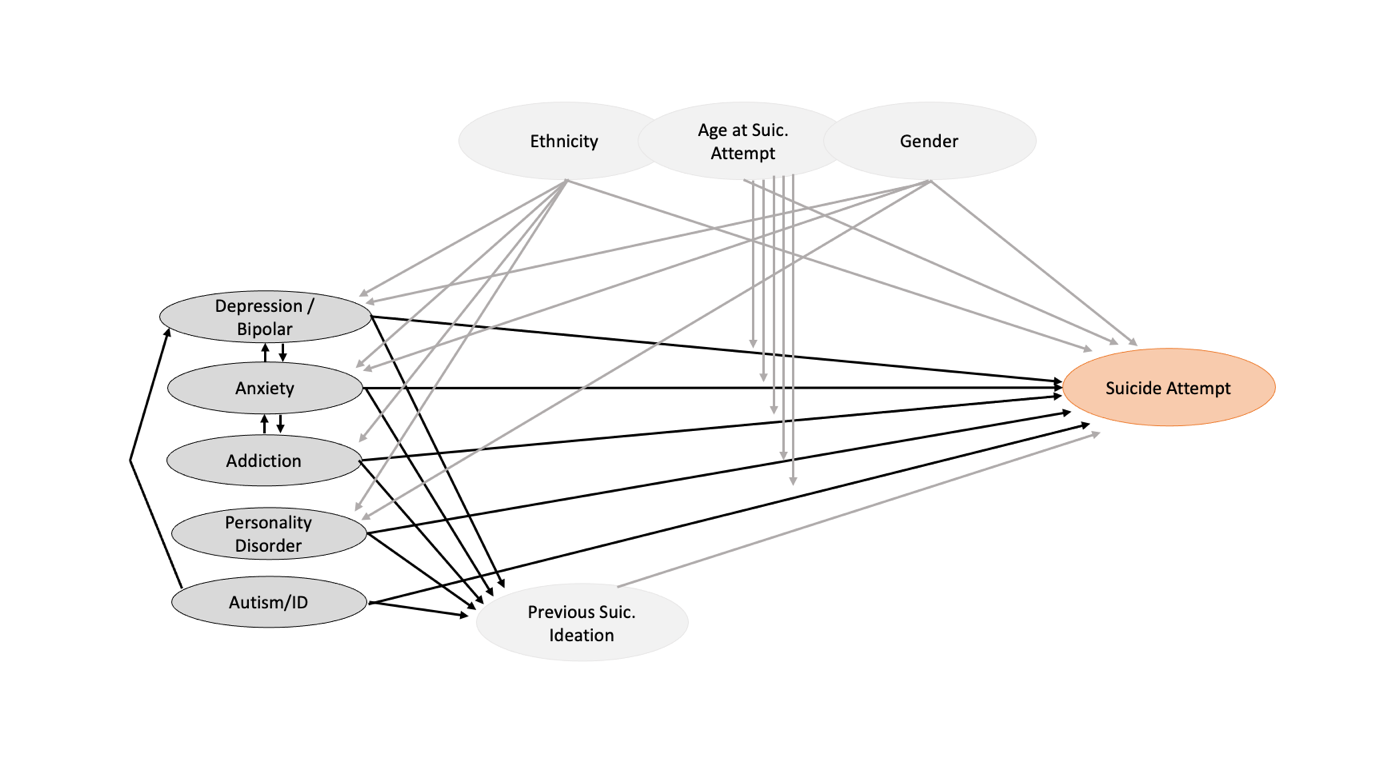

**Supplementary figure 4.** Causal diagram depicting psychiatric comorbidity variables in relation to Suicide Attempt. An arrow’s direction represents the theoretical direction of causal effect. An arrow pointing to the middle body of another arrow indicates a theoretical moderation effect. Dark grey = predictor variables. Light grey = control variables. Red = predicted variable.

#### Supplementary Tables

**Table 2 supplement.** Supplementary results from bias-reduced Logistic Regression analysis on **suicidal ideation in people with epilepsy.** Model fit statistics by target risk factor including the D3 likelihood ratio test for multiply imputed data and average R2 value across imputed data sets.

| **Term** | **D3** | **p** | **R2** |
| --- | --- | --- | --- |
| Age at SI | 7.5 (1, 1728711) | 0.006 | 0.005 |
| Age at SI (non-linear) | 8.3 (2, 2131450) | 0.000 | 0.011 |
| Gender: Female (vs Male) | 2.1 (1, 1873551) | 0.145 | 0.002 |
| Ethnicity: White, Asian, Black, Mixed, Other (vs White) | 0.1 (1, 2260) | 0.796 | 0.000 |
| Any neuropsychiatric comorbidities: Yes (vs No) | 5.7 (3, 84900) | 0.001 | 0.013 |
| - Depression / Bipolar: Yes (vs No) | 9.5 (5, 507087) | 0.000 | 0.032 |
| - Anxiety / stress related: Yes (vs No) | 3 (5, 488139) | 0.011 | 0.014 |
| - Substance misuse: Yes (vs No) | 3.8 (3, 70620) | 0.009 | 0.011 |
| - PDD / ID: Yes (vs No) | 45.8 (1, 1206033) | 0.000 | 0.029 |

**Table 3 supplement**. Supplementary results from bias-reduced Logistic Regression analysis on **suicide attempt-related hospitalisation in people with epilepsy.** Model fit statistics by target risk factor including the D3 likelihood ratio test for multiply imputed data and average R2 value across imputed data sets.

| **Term** | **D3** | **p** | **R2** |
| --- | --- | --- | --- |
| Age at SA | 2.8 (1, 248796) | 0.094 | 0.002 |
| Age at SA (Non-linear) | 9.4 (2, 318133) | 0.000 | 0.012 |
| Gender: Female (vs Male) | 4.7 (1, 255993) | 0.030 | 0.004 |
| Ethnicity: White, Asian, Black, Mixed, Other (vs White) | 7.4 (1, 1010) | 0.007 | 0.006 |
| Any neuropsychiatric comorbidities: Yes (vs No) | 15.6 (3, 45193) | 0.000 | 0.038 |
| - Depression / Bipolar: Yes (vs No) | 9.3 (5, 721896) | 0.000 | 0.033 |
| - Anxiety / stress related: Yes (vs No) | 3.1 (5, 513571) | 0.009 | 0.011 |
| - Substance misuse: Yes (vs No) | 3.5 (3, 44267) | 0.014 | 0.008 |
| - PDD / ID: Yes (vs No) | 33.8 (1, 202937) | 0.000 | 0.020 |
| Previous SI: Yes (vs No) | 11.9 (4, 161049) | 0.000 | 0.038 |

**Table 4 supplement**. Supplementary results from bias-reduced Logistic Regression analysis on **suicidal ideation in people with FDS.** Model fit statistics by target risk factor including the D3 likelihood ratio test for multiply imputed data and average R2 value across imputed data sets.

| **Term** | **D3** | **p** | **R2** |
| --- | --- | --- | --- |
| Age at SI | 56.3 (1, 3424225) | 0.000 | 0.054 |
| Age at SI (Non-linear) | 28.6 (3, 6658039) | 0.000 | 0.088 |
| Gender: Female (vs Male) | 1.7 (1, 2810711) | 0.191 | 0.002 |
| Ethnicity: White, Asian, Black, Mixed, Other (vs White) | 18 (1, 14485) | 0.000 | 0.020 |
| Any neuropsychiatric comorbidities: Yes (vs No) | 8.8 (3, 739768) | 0.000 | 0.028 |
| - Depression / Bipolar: Yes (vs No) | 5.6 (5, 7080170) | 0.000 | 0.030 |
| - Anxiety / stress related: Yes (vs No) | 5.7 (5, 6035580) | 0.000 | 0.031 |
| - Personality Disorder: Yes (vs No) | 8.7 (3, 696693) | 0.000 | 0.029 |

**Table 5 supplement**. Supplementary results from bias-reduced Logistic Regression analysis on **suicide attempt-related hospitalisation in people with FDS.** Model fit statistics by target risk factor including the D3 likelihood ratio test for multiply imputed data and average R2 value across imputed data sets.

| **Term** | **D3** | **p** | **R2** |
| --- | --- | --- | --- |
| Age at SA | 112.1 (1, 1350724) | 0.000 | 0.118 |
| Age at SA (Non-linear) | 45.2 (3, 1962553) | 0.000 | 0.157 |
| Gender: Female (vs Male) | 0 (1, 1019202) | 0.920 | 0.000 |
| Ethnicity: White, Asian, Black, Mixed, Other (vs White) | 6.3 (1, 130198) | 0.012 | 0.005 |
| Any neuropsychiatric comorbidities: Yes (vs No) | 6.9 (3, 13861242) | 0.000 | 0.017 |
| - Depression / Bipolar: Yes (vs No) | 3.4 (5, 2532484) | 0.004 | 0.013 |
| Previous SI: Yes (vs No) | 5.9 (4, 3672085) | 0.000 | 0.020 |
